# Prediction of Cardiovascular Markers and Diseases Using Retinal Fundus Images and Deep Learning: A Systematic Scoping Review

**DOI:** 10.1101/2024.04.17.24305957

**Authors:** Livie Yumeng Li, Anders Aasted Isaksen, Benjamin Lebiecka-Johansen, Kristian Funck, Vajira Thambawita, Stine Byberg, Tue Helms Andersen, Ole Norgaard, Adam Hulman

**Author notes:** Corresponding authors: Livie Yumeng Li, Palle Juul-Jensens Boulevard 11, Entrance A, Steno Diabetes Center Aarhus, Aarhus N, DK-8200, Denmark, Adam Hulman, Palle Juul-Jensens Boulevard 11, Entrance A, Steno Diabetes Center Aarhus, Aarhus N, DK-8200, Denmark.

## Abstract

**Background:** Cardiovascular risk prediction models based on sociodemographic factors and traditional clinical measurements have received significant attention. With rapid development in deep learning for image analysis in the last decade and the well-known association between micro- and macrovascular complications, some recent studies focused on the prediction of cardiovascular risk using retinal fundus images. The objective of this scoping review is to identify and describe studies using retinal fundus images and deep learning to predict cardiovascular risk markers and diseases.

**Methods:** We searched MEDLINE and Embase for peer-reviewed articles on 17 November 2023. Abstracts and relevant full-text articles were independently screened by two reviewers. We included studies that used deep learning for the analysis of retinal fundus images to predict cardiovascular risk markers (e.g. blood pressure, coronary artery calcification, intima-media thickness) or cardiovascular diseases (prevalent or incident). Studies that used only predefined characteristics of retinal fundus images (e.g. tortuosity, fractal dimension) were not considered. Study characteristics were extracted by the first author and verified by the senior author. Results are presented using descriptive statistics.

**Results:** We included 24 articles in the review, published between 2018 and 2023. Among these, 21 (88%) were cross-sectional studies and eight (33%) were follow-up studies with outcome of clinical CVD. Five studies included a combination of both designs. Most studies (n=23, 96%) used convolutional neural networks to process images. We found nine (38%) studies that incorporated clinical risk factors in the prediction and four (17%) that compared the results to commonly used clinical risk scores in a prospective setting. Three of these reported improved discriminative performance. External validation of models was rare (n=5, 21%). Only four (17%) studies made their code publicly available.

**Conclusions:** There is an increasing interest in using retinal fundus images in cardiovascular risk assessment. However, there is a need for more prospective studies, comparisons of results to clinical risk scores and models augmented with traditional risk factors. Moreover, more extensive code sharing is necessary to make findings reproducible and more impactful beyond a specific study.

## Introduction

Cardiovascular diseases (CVDs) are the leading causes of mortality globally.^1^ Clinical risk prediction models can help to identify individuals at high risk and target preventive efforts including lifestyle and pharmacological interventions.^2^ There are some well-established and validated CVD prediction models based on sociodemographic factors and traditional clinical variables in the general population and in selected subpopulations, like the Framingham score,^3^ SCORE2,^4^ and QRISK models.^5^ A recent comprehensive systematic review called for validating existing tools, tailoring existing models to specific populations, and identifying new data sources to be included in models, instead of testing small alterations of the established clinical risk models.^6^

Developments in machine learning for image analysis in the last decade have made it feasible to include images as a data type in risk prediction models, potentially in combination with traditional risk factors (multimodal prediction models). Retinal fundus imaging is a relatively simple and the only non-invasive method used for assessing the state of blood vessels in the body. People with type1 and type 2 diabetes are regularly invited to diabetic retinopathy screening with retinal fundus images, which makes it a potentially relevant predictor and allows us to follow the progression in microvascular disease. It’s likely that the underlying disease causes damage to all blood vessels, resulting in both micro- and macrovascular complications. The state of the blood vessels in the retina could reflect that of blood vessels elsewhere. There is a well-established association between diabetic retinopathy and both micro- and macrovascular complications, including CVD.^7,8^ However, it remains an open question whether these associations can be translated into clinically relevant predictors. Instead of using disease status as the predictor, the actual images might carry more relevant information for CVD prediction.

In a preliminary search, we identified a review on the topic of our scoping review.^9^ However, there are some limitations of this work. Firstly, the search procedure was described only very briefly. Secondly, the authors only considered open-access articles in the review.Thirdly, they had a focus on diabetic retinopathy grading and image segmentation unrelated to cardiovascular disease. Since the review was published, we have identified several relevant articles on the topic.^10,11^ Finally, we aim to evaluate the studies from a more clinical perspective (e.g. performance comparison with or added value to existing risk scores).

We chose to conduct a systematic scoping review instead of a classical systematic review because scoping reviews serve to scope a body of literature, examine research practices, and clarify concepts, which fit well with our goals.^12^ We aimed to describe studies, with an emphasis on clinical perspectives, that used retinal fundus images and deep learning for the prediction of cardiovascular markers and diseases.

## Methods

The scoping review was conducted in accordance with the Joanna Briggs Institute (JBI) methodology for scoping reviews and reported according to the Preferred Reporting Items for Systematic Reviews and Meta-analyses extension for scoping review (PRISMA-ScR) guidelines.^13^ The scoping review protocol was published on Figshare on December 19, 2023.^14^

### Eligibility Criteria

#### Participants

We considered studies analysing data from human participants regardless of their health status (e.g. population-based studies or cohorts with a specific condition).

#### Context

This scoping review focuses on studies in the clinical research context, regardless of the geographical location, ethnicity, and gender composition of the study populations. Methodological articles comparing different methods for CVD risk prediction based on retinal fundus images were considered if any of the methods used deep learning.

#### Types of Sources

We included peer-reviewed articles (e.g. original articles, brief reports; peer-reviewed full-length articles in conference proceedings). Included studies needed to be human clinical studies (including methodological studies with examples using clinically relevant outcomes and measures). Furthermore, we only included studies that used deep learning to predict cardiovascular markers, or presence of CVD, or CVD incidence based on retinal fundus images. Studies could also use a deep learning-derived score from retinal fundus images (not pre-defined retinal features), to predict cardiovascular markers or diseases.

We excluded review articles, editorials, conference abstracts and preprints. Studies were excluded if they only included optical coherence tomography (OCT) or other eye images for prediction, if they extracted pre-defined features of the vessel network (e.g. tortuosity, fractal dimension) and then associated them with CVD markers, or if they predicted factors that were not based on measurements of the cardiovascular system, but merely risk factors of CVD (e.g. age, sex, cholesterol, HbA1c). We also excluded association studies, even if they investigated the association between a deep learning-derived score and cardiovascular markers or diseases, since such studies do not report any predictive performance metrics.

Articles in languages not understood by the review team (English, Danish, Swedish, Norwegian, Hungarian, German, Polish, and Chinese) that were considered eligible by title and abstract were not included in the synthesis but would have been included in an appendix for others to analyse.

### Search Strategy

We considered MEDLINE and Embase as databases for the search. Most of the data science literature focuses on diabetic retinopathy grading, most likely due to the availability of open-access datasets from this domain. As this task is not directly relevant from a cardiovascular perspective and it is unlikely that such datasets have detailed phenotyping of cardiovascular health or follow-up for hard endpoints, we did not consider technical databases.

Information specialist THA conducted the search on November 17, 2023. The search comprised three key concepts: retina, cardiovascular diseases and artificial intelligence/machine learning. Each concept was searched using Medical Subject Headings (MeSH) and free-text words, and no limits were applied. The search string was developed in MEDLINE and subsequently translated to Embase. The search string was tested against eight key articles within the field and reviewed by another information specialist (ON). The full search string in both databases is available on Figshare.^15^

After the selection process, we used the software tool citationchaser^16^ to retrieve all references within and all articles citing the included articles and previous reviews within the same topic. Retrieved articles were screened to find all relevant articles.

### Study Selection

Following the search, all identified citations were collated and uploaded into EPPI-Reviewer 6 and duplicates were removed.^17^

In the screening phase, two independent reviewers screened titles and abstracts to assess eligibility. At the beginning, we had a pilot screening workshop examine the in- and exclusion of about 25 abstracts. In the official screening, the reviewers met to assess alignment in the process after screening 10% of the abstracts. After all titles and abstracts had been screened, full-text versions of relevant articles were retrieved and assessed in detail against the eligibility criteria by two or more independent reviewers. Reasons for the exclusion of articles at full-text screening were recorded and reported in the Results section. Any disagreements between the reviewers at any stage of the selection process were resolved through discussion. If there was no consensus after this, the senior author (AH) made the final decision. The study selection process is presented using a flow diagram.

### Data Extraction

Research questions (Table 1) were pre-specified and published in the scoping review protocol.^14^ Data was extracted from included articles by the first author and verified by the senior author. Data was collected using a data extraction instrument developed based on the research questions. The extracted data included specific details about the study methods and characteristics relevant to the review questions listed in the protocol (e.g. first author and year of publication; study population and design; CVD outcomes; deep learning model used; predictive performance; comparison to clinical risk scores if included). The identified studies described in the data extraction table are published on Figshare.^15^

**Table 1.** A priori defined research questions.

|  |  |
| --- | --- |
| 1 | To what extent is deep learning used for predicting cardiovascular risk using retinal fundus images and what characterises the studies? |
| 2 | Which cardiovascular markers and diseases are predicted in the studies? |
| 3 | What are the study populations (e.g. general population, type 2 diabetes)? |
| 4 | How diverse are study populations (age, sex, ethnicity, geographic location)? |
| 5 | How do prediction models perform? Which clinical prediction performance metrics are used? |
| 6 | Are deep learning models compared to classical clinical risk prediction models? |
| 7 | Are the models validated internally or externally? |
| 8 | Are the models tested or implemented in a clinical setting? |
| 9 | What kind of deep learning architectures are used (e.g. convolutional neural networks)? |
| 10 | In the case of incident CVD as the outcome (follow-up studies), how is the time-to-event nature of data handled? |
| 11 | Are there studies combining other data modalities (e.g. tabular/structured clinical data) with retinal fundus images? |
| 12 | Are there any publicly available datasets (open / restricted by payment / private)? |
| 13 | Are the models and the code publicly available (e.g. on Hugging Face or GitHub)? |

### Data Analysis

Study characteristics are aggregated using descriptive statistics (frequencies and percentages). A narrative summary accompanies the tabulated and charted results and describes how the results relate to the review’s objective and questions.

## Results

The search resulted in 1,990 records, of which 172 were duplicates (Fig 1). After screening the titles and abstracts of the remaining 1,818 records, 1,790 were excluded as irrelevant. Of the 28 records included for full-text screening, 10 were excluded because they did not use deep learning (n=5), were not original articles (n=2), did not focus on CVD (n=2) or did not use fundus retinal images (n=1). Another six records were identified in citations and references of the included studies and previous reviews on similar topics.^9,18^ In total, 24 studies were included in this scoping review, all of which were published after 2018.

**Fig 1.**
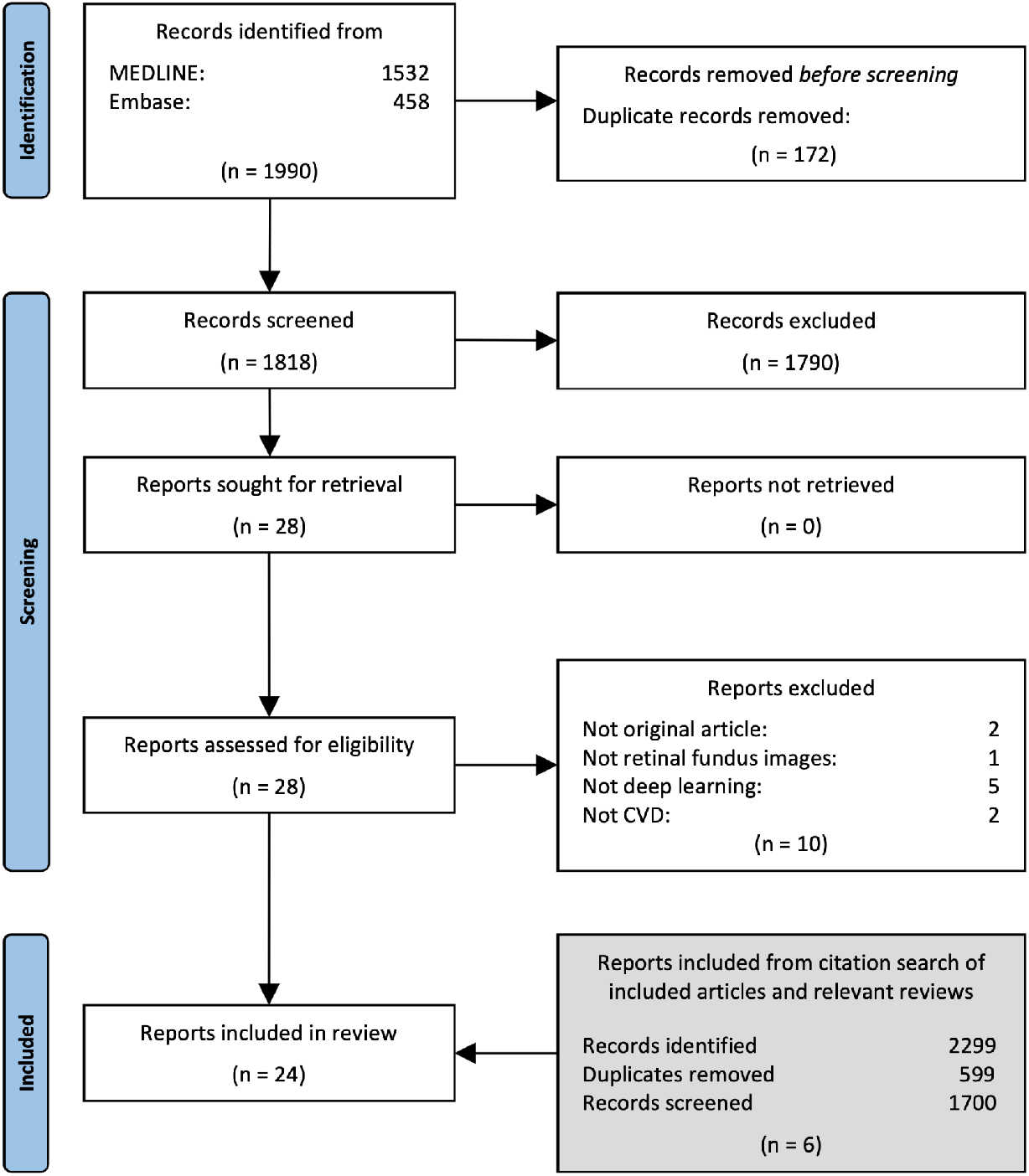
PRISMA flow chart

Among the identified studies, 11 out of 24 (46%) predicted markers of subclinical CVD as the outcome, 18 predicted clinical CVD, and five studies included both types of outcomes. Ten out of 18 studies were cross-sectional using retinal fundus images to predict prevalent CVD, and eight were cohort studies including clinical CVD.

Markers of subclinical CVD included systolic and diastolic blood pressure, left ventricular characteristics, brachial-ankle pulse-wave velocity, coronary artery calcium (CAC) score, and carotid intima-media thickness (Table 2). Clinical CVD included hypertension, peripheral arterial disease, coronary heart disease, cerebrovascular disease, stroke, myocardial infarction, heart failure, and CVD mortality. Apart from the desired outcomes listed in the studies, one study had an outcome described as CVD without further details, and one study had the outcome described as major adverse cardiovascular events (MACE) without further details included.

**Table 2.** Study design and outcome of included studies.

|  | **Markers of subclinical CVD** | **Clinical CVD** |
| --- | --- | --- |
| **Cross-sectional studies** | systolic and diastolic blood pressure<sup>20,32,34,42</sup><br>left ventricular characteristics<sup>43</sup><br>brachial-ankle pulse-wave velocity<sup>44</sup><br>coronary artery calcium score<sup>28–30,45–47</sup><br>carotid intima-media thickness<sup>33</sup> | hypertension<sup>48,49</sup><br>peripheral arterial disease<sup>50</sup><br>coronary heart disease<sup>10,51</sup><br>cerebrovascular disease<sup>10</sup><br>stroke<sup>19,52–54</sup><br>diagnosis of CVD<sup>55</sup> |
| **Follow-up studies** |  | major adverse cardiovascular events<sup>32</sup><br>peripheral arterial disease<sup>20</sup><br>coronary heart disease<sup>20,29</sup><br>stroke<sup>20,21,29,30</sup><br>myocardial infarction<sup>20,21,30,43</sup><br>major heart failure<sup>21,30</sup><br>CVD mortality<sup>28–30,33</sup> |

Most studies were conducted in the general population or without the mention of specific patient groups (22 out of 24). Two studies had more specific study populations, one included people with atrial fibrillation,^19^ and another included people with type 1 diabetes or type 2 diabetes.^20^ We used the first authors’ first affiliation as a proxy for the geographical location of the studies. More than half of the identified studies were conducted in Asia (13 out of 24, 54%): five from South Korea, four from China, and four from Singapore. We did not identify any studies from Africa, South America or Australia.

Most studies (n=23, 96%) used convolutional neural networks to process images. One study used the vision transformer deep learning architecture.^21^ No studies used a deep learning framework specifically developed for time-to-event data. Several performance metrics were used in the studies. The most often used predictive performance metric was discrimination, characterised by the area under the receiver operating characteristic curve (AUROC), also referred to as C-statistic or concordance index in time-to-event settings (n=18 out 24, 75%). For continuous outcomes, the most often used metrics were accuracy (n=6, 25%), mean absolute error (MAE) and the coefficient of determination also known as r-squared (n=3).

We found nine studies that combined clinical risk factors (in tabular data form) with images as predictors in their studies. Five out of nine included clinical risk factors in the deep learning models, whereas the other four applied a two-step approach extracting new variables using deep learning and then including them in Cox regression or Poisson regression models. Five studies compared the performance with established clinical risk scores such as Framingham Risk Score,^22^ Systematic Coronary Risk Assessment (SCORE),^23^ QRISK3,^24^ and the Pooled Cohort Equation for atherosclerotic CVD (PCE-ASCVD) (Table 3).^25^ One study presented a comparison with a customised clinical risk prediction model developed as part of the same study. Four studies reported incremental improvement in the predictive performance after adding retinal fundus images or scores derived from them to the clinical risk score or model of their choice.

**Table 3.**
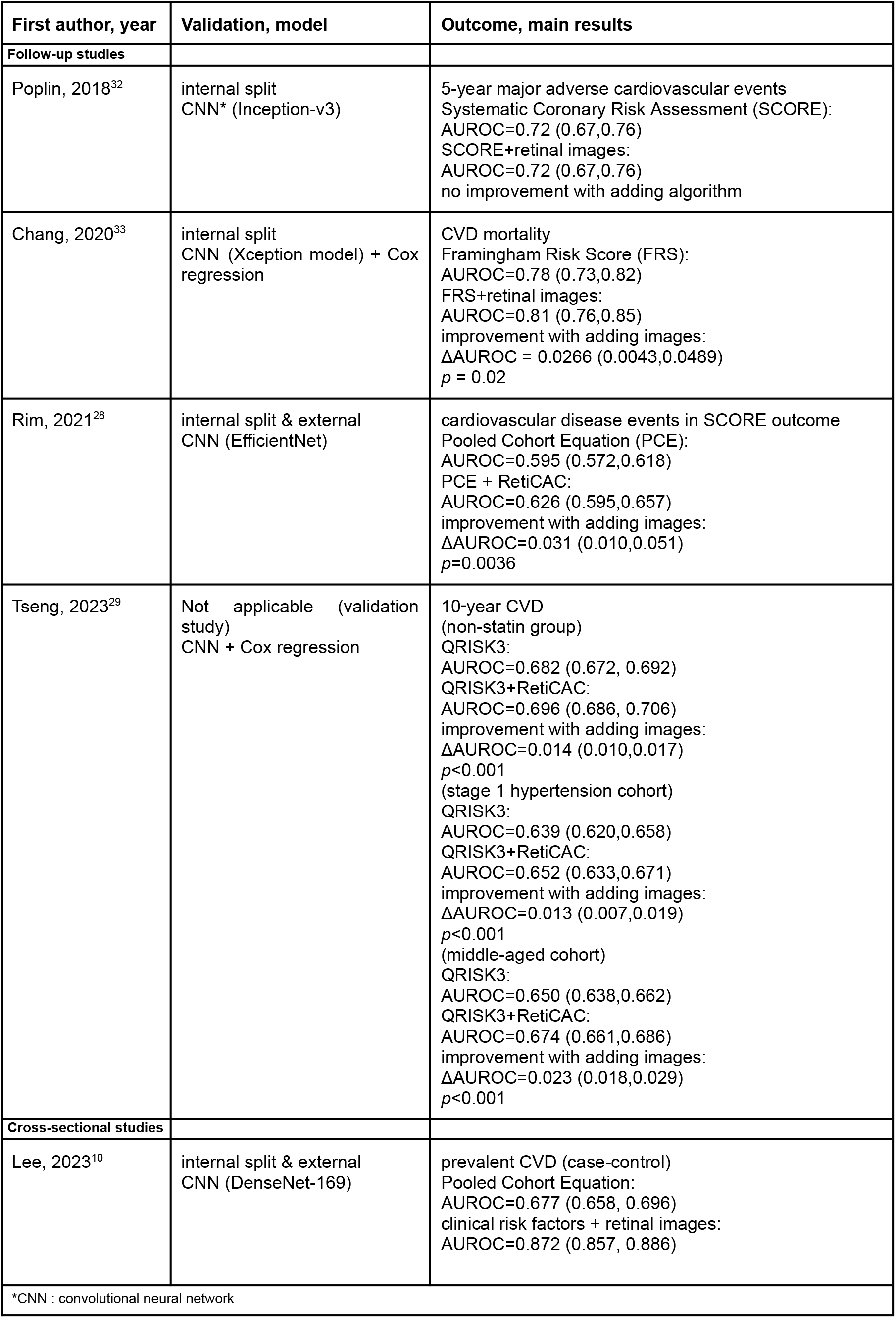
Characteristics of studies that compared retina-based prediction models with established clinical cardiovascular risk scores.

| First author, year | Validation, model | Outcome, main results |
| --- | --- | --- |
| **Follow-up studies** |  |  |
| Poplin, 2018<sup>32</sup> | internal split<br>CNN* (Inception-v3) | 5-year major adverse cardiovascular events<br>Systematic Coronary Risk Assessment (SCORE):<br>AUROC=0.72 (0.67,0.76)<br>SCORE+retinal images:<br>AUROC=0.72 (0.67,0.76)<br>no improvement with adding algorithm |
| Chang, 2020<sup>33</sup> | internal split<br>CNN (Xception model) + Cox<br>regression | CVD mortality<br>Framingham Risk Score (FRS):<br>AUROC=0.78 (0.73,0.82)<br>FRS+retinal images:<br>AUROC=0.81 (0.76,0.85)<br>improvement with adding images:<br>ΔAUROC = 0.0266 (0.0043,0.0489)<br>$p = 0.02$ |
| Rim, 2021<sup>28</sup> | internal split & external<br>CNN (EfficientNet) | cardiovascular disease events in SCORE outcome<br>Pooled Cohort Equation (PCE):<br>AUROC=0.595 (0.572,0.618)<br>PCE + RetiCAC:<br>AUROC=0.626 (0.595,0.657)<br>improvement with adding images:<br>ΔAUROC=0.031 (0.010,0.051)<br>$p=0.0036$ |
| Tseng, 2023<sup>29</sup> | Not applicable (validation<br>study)<br>CNN + Cox regression | 10-year CVD<br>(non-statin group)<br>QRISK3:<br>AUROC=0.682 (0.672, 0.692)<br>QRISK3+RetiCAC:<br>AUROC=0.696 (0.686, 0.706)<br>improvement with adding images:<br>ΔAUROC=0.014 (0.010,0.017)<br>$p<0.001$<br>(stage 1 hypertension cohort)<br>QRISK3:<br>AUROC=0.639 (0.620,0.658)<br>QRISK3+RetiCAC:<br>AUROC=0.652 (0.633,0.671)<br>improvement with adding images:<br>ΔAUROC=0.013 (0.007,0.019)<br>$p<0.001$<br>(middle-aged cohort)<br>QRISK3:<br>AUROC=0.650 (0.638,0.662)<br>QRISK3+RetiCAC:<br>AUROC=0.674 (0.661,0.686)<br>improvement with adding images:<br>ΔAUROC=0.023 (0.018,0.029)<br>$p<0.001$ |
| **Cross-sectional studies** |  |  |
| Lee, 2023<sup>10</sup> | internal split & external<br>CNN (DenseNet-169) | prevalent CVD (case-control)<br>Pooled Cohort Equation:<br>AUROC=0.677 (0.658, 0.696)<br>clinical risk factors + retinal images:<br>AUROC=0.872 (0.857, 0.886) |
| *CNN : convolutional neural network |  |  |

External validation of models was rare (n=5 out of 24, 21%), while internal validation was a common practice. Eight studies used cross-validation, and 12 split and set part of their study sample aside for validation. No study used a fully open-access dataset. Eight studies used data from the UK Biobank,^26^ which is available to anyone for a data access fee. Only four studies made their code publicly available on GitHub. None of the studies reported randomised clinical trials to evaluate the implementation of the developed algorithms and scores.

## Discussion

We identified and described studies exploring the integration of retinal fundus images in the prediction of cardiovascular markers and diseases. The majority of these studies were cross-sectional, lacking examination of predictive utility for hard clinical endpoints in prospective settings. Few studies compared their models with established cardiovascular risk scores, evaluating the potential of the multimodal approaches. In most of these cases, including retina images led to small improvements in discriminative performance. However, other clinically relevant metrics, like calibration, were often overlooked.

Clinical risk prediction models are traditionally developed using regression-based statistical methods that can only handle categorical and numerical variables i.e. tabular data. Recent advancements in deep learning allowed the analysis and integration of images with promising results in various medical domains, including clinical risk assessment of cardiovascular health.^27^ A multimodal approach offers the opportunity to extract novel insights from new, and often routinely collected data types, expanding the utilisation of available clinical data.

We found a series of studies on prediction of CAC from one research group, where they developed and validated a retinal fundus image-based score referred to as Reti-CVD or RetiCAC, and its utility was compared to existing CVD risk scores.^28–30^ The score was proposed as a non-invasive and cost-efficient alternative to the computer tomography-derived CAC score. Multiple validation studies investigated the incremental predictive value of RetiCAC on top of established clinical risk prediction models. Rim et al. added the score to PCE-ASCVD to predict SCORE CVD events and Tseng et al. added the score to QRISK3 to predict 10-year CVD respectively.^28,29^ They both reported small improvements in predictive performance after adding retinal image-derived scores to the models. In another study, Reti-CVD was used to identify people at intermediate and high risk of CVD according to established clinical risk scores (PCE-ASCVD, QRISK3 and Framingham Risk Score), and reported that it could effectively identify these groups.^11^ Lee et al. conducted a regulated pivotal trial (single-centre conformity design, confirmatory retrospective analysis) to validate the efficacy of Reti-CVD for stratification of CVD risk.^30^ The trial concluded superior performance in risk stratification compared to some subclinical CVD markers (carotid intima media thickness, pulse wave velocity), and non-inferiority to CAC score-based risk stratification. The commercialised product based on the RetiCAC/Reti-CVD score obtained CE approval as a class IIa medical device in the European Union and several countries in Asia.^31^ However, our search has not identified any randomised controlled trial conducted to evaluate the actual clinical impact of using the score.

In addition to the studies focusing on CAC prediction, three other studies compared their predictive performance with established clinical risk scores. Poplin et al. used a deep learning model to predict the 5-year MACE from retinal images.^32^ They compared the predictive performance of the original SCORE model to its image-augmented version They did not find evidence for improved predictive performance by including retinal images. Chang et al. had a similar approach to the outcome of CVD mortality using the Framingham Risk Score. This study reported minor improvements by integrating images into the model.^33^ In a case-control study, Lee et al. predicted the prevalence of CVD using a multimodal deep learning model integrating retinal images and traditional risk factors and compared the results to the predictive ability of PCE-ASCVD.^10^ They found major performance improvement by using the deep learning model.

When examining performance evaluations, we found a strong focus on model discrimination, which describes how well a model ranks predicted probabilities between individuals with and without an event. However, in a clinical setting, CVD risk scores are often used with a specific threshold (e.g. 10%) to make decisions on interventions, e.g. treatment intensification. Therefore, good discrimination alone is not sufficient, but calibration is highly relevant as well, which was overlooked by most of the studies. A miscalibrated model can lead to resource misallocation. Individuals incorrectly predicted with a high risk of CVD may be administered unnecessary tests or treatments, while those who could benefit from interventions may be overlooked.

Apart from the use of clinically relevant performance metrics for model evaluation, fair comparisons between existing and new models are also of high importance. Some of the identified studies reported that retinal images can be used to predict demographic factors (e.g. age, sex) and traditional clinical risk factors (e.g. blood pressure, body mass index).^32^ Many of these factors are associated with CVD risk, therefore, good performance of a prediction model based only on images does not necessarily mean that retinal images have additional value for CVD risk prediction, which is the underlying motivation of most identified studies. Models might achieve good performance by indirectly predicting risk factors that are easier to collect than retinal images. ^32^ This question can be partly addressed by stratifying participants by clinical risk factors possible to predict based on retinal images (e.g. age, sex).^34^ Another solution is to conduct incremental comparisons integrating images with traditional clinical risk factors, however, one should be careful with interpreting the results if important, unobserved risk factors are not included in the models.

Although studies demonstrated the potential of deep learning to contribute to improvements in cardiovascular risk assessment, methodological challenges, like insufficient validation, and poor reproducibility due to the lack of openness about data and code, must be addressed by future studies. Moreover, diverse study populations, both for development and validation, would improve the equality and fairness of potential clinical applications.^35,36^ No impact study was identified in our scoping review, which is a crucial missing link to measure the actual clinical utility of retinal image-based prediction models in CVD risk assessment.

Although most deep learning models were originally not designed to analyse time-to-event data, recent developments enabled them to account for this special data structure.^37,38^ However, none of the identified studies applied such a time-to-event framework. The most common approach to circumvent this limitation was a two-step approach combining deep learning with classical survival analysis methods.^20,28–30^ In step one, the deep learning model takes one or a pair of retinal fundus images as input and outputs an intermediate score or abstract feature representation. In step two, the score is used, often in combination with clinical risk factors, as input for a Cox or Poisson regression model, that then outputs predicted probabilities. Since these statistical models are consistent with what is used in classical survival analyses, the comparison and validation are analogous to established risk scores and risk prediction models. Some studies ignored the time-to-event nature of data, which can lead to underestimation of CVD risk especially in high-risk groups.^39^

### Strengths and Limitations

As a systematic scoping review, the strength of our study is the comprehensive and systematic exploration of the scope. The search strategy was thoroughly developed and documented contributing to transparency and reproducibility. Our review was written with a broad target audience addressing study characteristics relevant from both a clinical and a data science perspective. One limitation of our study is that we searched only medical databases, and might have overlooked studies from the technical sciences and engineering communities. To address this, we screened all references and citing articles of the included studies and reviews similar to ours. This process resulted in six additional articles, five of which were not indexed in the medical databases we searched.

### Perspectives

In this scoping review, we identified a gap between the proposed methods in clinical research and the latest developments in the field of deep learning. A fully deep learning-driven multimodal time-to-event prediction model could bring new insight to cardiovascular disease risk analysis and risk progression by learning from a series of images in longitudinal settings. Our findings support that improvements are needed in the recently proposed five critical quality criteria for artificial intelligence-based prediction model development and validation studies.^40^ Research in this highly interdisciplinary field must strive to report adequate details from both the clinical and data science aspects. Further collaboration and better communication between the two fields are crucial to raising the quality of studies. More validation studies with a focus on diversity regarding geographical location and ethnicity should be conducted in the future. At the same time, efforts should be spent to promote open-access datasets and algorithms to align with the FAIR principles.^41^ With the new AI Act approved in the European Parliament and similar proposals introduced globally, it is expected that research utilising AI technology will be forced to adhere to stricter regulations regarding transparency, accountability, and ethical considerations throughout the developmental and implementation lifecycle.

## Conclusion

Technological advancements in the field of artificial intelligence offer the potential to integrate new data types in analyses that were traditionally based on tabular data. Our scoping review presented the landscape of how deep learning methods integrate retinal fundus images in cardiovascular risk prediction. Some evidence shows improvements in predictive performance when adding images to clinical risk factors, but similar to prediction research in general, methodological weaknesses have to be overcome before the full potential can be exploited. There is a need for conducting more prospective studies, promoting performance metrics other than discrimination, increasing efforts on external validation of findings in datasets from diverse settings, and initiating impact studies to measure the clinical value of new multimodal risk assessment tools while improving the reproducibility of research. Working with images and deep learning models requires new methodological skills and close collaboration between the clinical research and data science communities so that developed models are described and tested adequately, and most importantly that they fulfil a validated clinical purpose.

## Author Contributions

LYL, AAI, BLJ, THA, ON, and AH conceptualised the study. THA and ON developed the search strategy with feedback from LYL, AAI, BLJ and AH. THA conducted the search. LYL, AAI, BLJ and AH screened abstracts. LYL and AH screened full-text articles. LYL extracted data from the identified articles. The data extraction forms were verified by AH. LYL analysed the data and presented the results. All authors contributed to the interpretation of the results. LYL wrote the original draft of the manuscript with support from AH. All authors read, edited and approved the final version of the manuscript. AH was responsible for the supervision of the project and funding acquisition.

## Acknowledgements

The authors are grateful to Helene Bei Thomsen (Steno Diabetes Center Aarhus) for her valuable comments on the Discussion.

## Data Availability

The study protocol and the completed data extraction forms were published elsewhere (see Methods section).

